# Nasopharyngeal Colonization, Associated Factors and Antimicrobial Resistance of *Streptococcus pneumoniae* among Children under 5 Years of Age in the Southwestern Colombia

**DOI:** 10.1101/2020.04.09.20058529

**Authors:** Gustavo Gámez, Juan Pablo Rojas, Santiago Cardona, Juan David Castillo, María Alejandra Palacio, Luis Fernando Mejía, José Luis Torres, Jaime Contreras, Laura Mery Muñoz, Javier Criales, Luis Felipe Vélez, Angélica María Forero, Yulieth Alexandra Zúñiga, María Eugenia Cuastumal, Leidy Johanna Acevedo, Álvaro de Jesús Molina, Johan Alexis Bolivar, Alejandro Gómez-Mejia, Jessica Lorena Morales, Sven Hammerschmidt

## Abstract

*Streptococcus pneumoniae* diseases are important causes of children death worldwide. Nasopharyngeal carriage of this pathobiont promotes bacterial spread and infections in the community. Here, a cross-sectional surveillance study was done to determine the proportion of nasopharyngeal colonization, antimicrobial susceptibility profile and associated factors in pediatric outpatients (southwestern Colombia, 2019). Data on factors associated with pneumococcal nasopharyngeal carriage were obtained through survey-based interviews. Nasopharyngeal swabs were collected and bacteria were microbiologically characterized. Antimicrobial susceptibility tests were done by VITEK-2. A logistic regression analysis was performed to examine associated factors. Tests with a *p*-value <0.05 were considered statistically significant. 452 children from the southwestern Colombia were examined and 41.8% carried *S. pneumoniae*. A higher pneumococcal carriage frequency was observed among participants <2-years and in individuals belonging to indigenous communities, which were not immunized against pneumococcus, because of lacking established immunization schemes. Additionally, children attending child-care institutions were also highly colonized by pneumococci. *S. pneumoniae* showed 57.7% non-susceptibility to benzyl-penicillin (meningitis-cut); 45.5% intermediate-sensitivity to benzyl-penicillin (oral-cut) and 21.7% to cefotaxime; and resistance to erythromycin (40.7%), tetracycline (36.0%), trimethoprim/sulfamethoxazole (24.9%), clindamycin (24.3%) and ceftriaxone (27.0%). The proportion of 41.8% of participants carrying *S. pneumoniae* shows a scenario with the presence of strains resistant to different antimicrobial agents (MDR and XDR), which constitutes important reservoirs of bacterial transmission by children <5-years in the southwest of Colombia. This situation could potentially lead to an onset of pneumococcal diseases. Hence, the need to expand conjugate pneumococcal immunization in the community and ensure compliance with established immunization schedules.

## Introduction

*Streptococcus pneumoniae*, also known as the pneumococcus, is a Gram-positive bacteria and a normal inhabitant of the nasopharyngeal microbiota of healthy children under 5 years old. The colonization process begins shortly after birth. Although pneumococcal carriage is usually asymptomatic, it can later serve as a reservoir for infections in children, the elderly, immunocompromised people and individuals with underlying diseases. However, *S. pneumoniae* is also a severe pathogen capable of causing diseases including community-acquired pneumonia (CAP), bacteremia, sepsis, meningitis, otitis media and sinusitis (1,2). It is estimated that diseases caused by this bacterium are a major public health problem worldwide, due to high morbidity and mortality rates (3). *S. pneumoniae* is the leading cause of lower respiratory tract infections worldwide, contributing to more deaths than all other etiologies combined (4). Nearly one million children under the age of 5 die each year due to diseases caused by the pneumococcus (2).

CAP constitutes a significant proportion of hospital admissions and the global burden of disease in children, being responsible for high mortality rates in infants, mainly in countries with low and medium incomes (5–9). *S. pneumoniae* is the leading cause of bacterial pneumonia, being identified in most cases, and is therefore considered a silent killer of children under 5 years of age (10,11). In Colombia, pneumonia is one of the main causes of mortality with 13 cases 100,000 deaths, with *S. pneumoniae* being its main etiological agent with a mortality rate of 3% (12). Likewise, for *S. pneumoniae* in Colombia, the average incidence is 0.28 cases per / 100,000 inhabitants, with a lethality between 13% and 27%, even with the appropriate treatment of individuals affected (13,14).

The asymptomatic carriage of *S. pneumoniae* has been identified as a prerequisite for the development of invasive and non-invasive diseases, and the carriers serve as sources of transmission of *S. pneumoniae* to other individuals in the community and within hospitals (15–18). Several clinical and demographic characteristics have been positively associated with an increase in the colonization of *S. pneumoniae*, such as infancy, overcrowding, childcare assistance, family size, sibling numbers, poverty, smoking and recent use of antibiotics (19–21). Although nasopharyngeal isolates are not useful for predicting the causative agent of invasive disease in individuals, they do reflect the epidemiological aspects of diseases caused by *S. pneumoniae* in the community (22,23). Bacteria inhabiting the upper respiratory tract of healthy children reflect the strains causing infection that are currently circulating in the community (24). Studies in the recent decades have gradually revealed the connection between pneumococcal carriage and invasive, and mucous infections caused by this pathobiont (15,18,25).

For many years, antibiotics such as penicillin and chloramphenicol have been used for the treatment of pneumococcal disease in low-and middle-income countries, given its effectiveness and low costs. Unfortunately, the dramatic increase in resistance to these and other antimicrobial agents worldwide has made the choice of antimicrobial drugs for *S. pneumoniae* infections increasingly difficult and expensive (28–31). Currently, prevention campaigns against pneumococcal infections are carried out using pneumococcal-conjugated vaccines (PCVs), which use the capsular polysaccharides of those specific serotypes most frequently associated with invasive pneumococcal diseases (IPDs) (1,26,27). In Colombia, the 7-valent pneumococcal conjugate vaccine was introduced in 2006 in the National Immunization Program, and replaced in 2011 by the 10-valent (2+1 doses).

The decrease in several primary risk factors, the implementation of better immunization strategies, and advances in the treatment of pneumococcal infections have made a substantial progress in recent years in reducing the burden of pneumococcal diseases. However, this has not been the same in all geographic regions of the world and more research and intervention efforts are still needed. In addition, nasopharyngeal colonization by antibiotic-resistant *S. pneumoniae* has been steadily increasing, representing potential dangers for the community (4,32,33). In Colombia, and particularly in the southwestern region (Departments of Valle del Cauca, Cauca, Putumayo and Nariño) (Figure 1), epidemiological data on *S. pneumoniae* are extremely limited. Therefore, the objective of this study was to determine the proportion of nasopharyngeal colonization, the profile of antimicrobial susceptibility and the possible associated factors in pediatric outpatients attending the Club Noel Children’s Clinical Foundation. This information will be useful for the implementation of more rational therapeutic and preventive strategies against pneumococcus in Southwestern Colombia, where the therapy for pneumococcal disease remains empirical due to the lack of rapid, sensitive and specific diagnostic tests.

**Figure 1.**
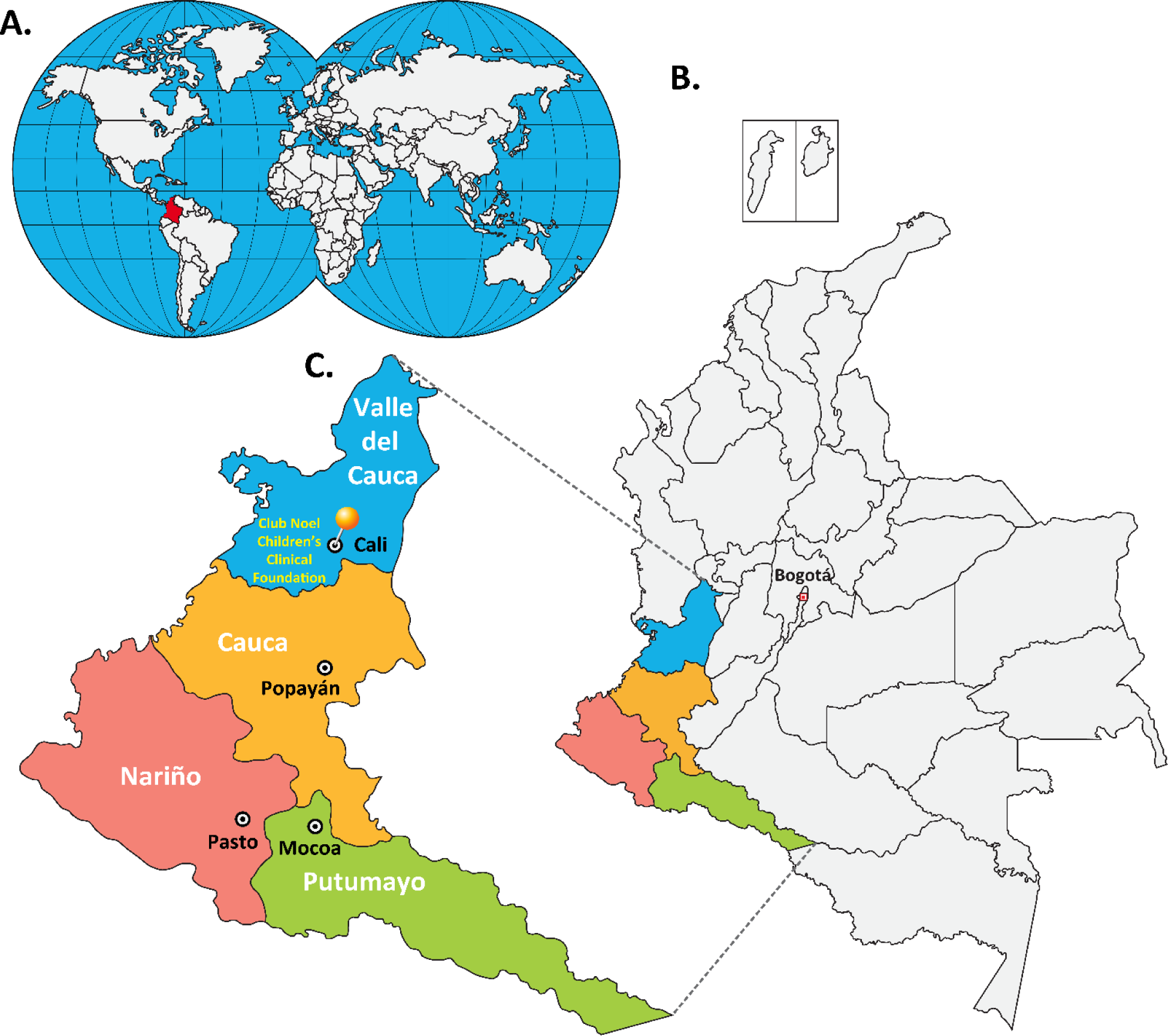
Geographic location of southwestern Colombia in the World. **A**. Republic of Colombia in the world. **B**. Southwestern region in the Colombian territory. **C**. Departments of Valle del Cauca, Cauca, Nariño and Putumayo and their capitals Cali, Popayán, Pasto and Mocoa, respectively. Club Noel Children’s Clinical Foundation is located in Santiago de Cali (Valle del Cauca). Bogotá is the capital of Colombia.

## 2. Methodology

### 2.1. Research Context, Population Definition and Inclusion Criteria

This work consisted on a cross-sectional study conducted for the southwestern region of Colombia (Figure 1), where 452 children under 5 years of age were randomly selected and prospectively involved in the surveillance in 2019. For convenience and logistical support, all children under five years of age eligible to participate in the study were those who attended the Club Noel Children’s Clinical Foundation of Cali (the main populated center of the Colombian southwest) and who were evaluated by external consult for pediatric control visits (34–37). The Club Noel Children’s Clinical Foundation is a second level pediatric hospital that operates every day of the year, serving up to 300 patients per day. Routine functions of the outpatient department include monitoring of children’s growth, immunization, nutritional counseling and management of ailments. Although the population attending in Club Noel Children’s Clinical Foundation comes mainly from the city of Cali, another large number of pediatric patients come from other nearby municipalities and departments, causing the study population to become a mixture of people from different races, and representatives of both urban and rural areas.

### 2.2. Sampling of the Study Population and Exclusion Criteria

The study population of this work was constituted by the pneumococcal colonizing isolates from the 452 children below 5 years old from the southwest Colombia involved in the investigation. This sample size was estimated using the general formula for a proportion of a single population with the following assumptions: 1) A total population of children under 5 years of age in the Colombian southwest of 734,372 (according to DANE projections as of June 30, 2018, the total population of the Colombian southwest would correspond to about 16% of the total national territory (7,981,162 of 49,879,349), of which about 669,400 would be children under 5 years of age) (38,39); 2) A prevalence rate of nasopharyngeal colonization by pneumococci of 50% (according to unpublished studies by our research group in the city of Medellin: 55.5%); 3) A 95% confidence level; AND 4) A marginal error or possible loss of information of 20%. The study participants were randomly associated and involved prospectively until the sample size was completed. The main exclusion criteria were: 1) Children under 5 years old with invasive pneumococcal disease diagnosed or critically affected by other diseases such as bronchopulmonary dysplasia, chronic immunodeficiency, cancer or any other acute, moderate or severe illness; 2) Children who had received antibiotics in the previous month or who had previously received any immunosuppressive medication such as prednisolone, cyclosporine, methotrexate, cyclophosphamide, azathioprine, mycophenolate mofetil or 5-fluorouracil; and 3) Those children whose parents or guardians were unwilling or unable to give their informed consent freely and spontaneously.

### 2.3. Collection of Clinical and Sociodemographic Information

This minimum risk level study was reviewed and approved by the Bioethics Committee of the Club Noel Children’s Clinical Foundation in Cali - Colombia. Initially, the study was socialized with parents or guardians of each child, who subsequently accepted the voluntary participation of their children by signing a written informed consent. Each participant was assigned a code and only the personnel involved in the investigation had confidential access to the information in the individual records. Then, the sociodemographic and housing data of the children under 5 years of age participating in the study were collected through a standardized and previously tested survey, applied to the parents or guardians of each child. The questionnaire of survey was mainly made up of closed questions that inquired about the date of birth, age, gender, race, conditions and socioeconomic status of the home, the constitution of the family nucleus, among others. Likewise, data were collected on the clinical history of the participating children such as nutritional status, breastfeeding, consumption of antibiotics and other medications, the history of the diseases suffered, and immunization records grouped into the following categories: not immunized, incomplete vaccination, vaccination in process (for children <1 year of age), and complete vaccination (for children> 1 year of age).

### 2.4. Nasopharyngeal Swab and Preservation of Biological Material

Nasopharyngeal swab samples were collected from each child selected to participate in the study using sterile flexible swabs (Copan, Brescia, Italy), according to standardized procedures. In brief, the swab was introduced through a nostril to the nasopharynx, where it was turned 180 degrees, and then carefully removed. Each swab collected was immediately introduced into a cryovial with 1 mL of STGG transport medium (skim milk solution-tryptone-glucose-glycerol) (40,41), for preservation at −30°C in the Microbiology Laboratory of Club Noel Children’s Clinical Foundation. Samples collected each week were transported on dry ice to the Central Research Laboratory of the School of Microbiology of the University of Antioquia in Medellín, Colombia.

### 2.5 Isolation, Cultivation, Identification and Cryopreservation of Pneumococci

To isolate and cultivate the pneumococcus after arrival of the material to the laboratory, a 10 µL aliquot of the sample was used for growth in tryptic soy agar (TSA) solid medium, supplemented with defibrinated sheep blood (5%), yeast extract (0.5%) and gentamicin (5 µg/mL). Pneumococci were incubated at 37°C and CO_2_ (5%) for a maximum of 24 hours, after which the following tests and assays were necessary to confirm their identification: 1) Evidence of Hemolysis (although the pneumococci are catalase-negative bacteria, as α-hemolytic microorganisms they can cause partial hemolysis due to the oxidation of iron in hemoglobin, which causes greenish colonies to occur when grown on blood agar plates), 2) Gram staining (pneumococci are Gram-positive), 3) Optochin sensitivity test (unlike other streptococci and α-hemolytic microorganisms, pneumococci are sensitive to 5 mg ethylhydrocupreine hydrochloride / optochin. Isolates with an inhibition zone ≥14 mm in diameter were considered susceptible to optochin), and 4) Bile solubility test (the pneumococcus is soluble in bile). The addition of small amounts of bile salts (2% sodium deoxycholate) results in complete destruction of the pneumococcus after a short incubation period. Finally, stocks were generated for the storage of the isolates, through the use of Todd-Hewitt supplemented with yeast extract (THY) liquid culture medium, supplemented with glycerol (25%) in cryovials, which were then preserved in an ultra-freezer at −80°C (2,41).

### 2.6. Determination of Susceptibility / Antibiotic Resistance (VITEK-2 System)

To test the susceptibility or resistance of the colonizing isolates against different antibiotics, fresh cultures of *S. pneumoniae* were made on Mueller-Hinton Agar plates, supplemented with Ram’s Blood (5%) and incubated for 14 hours at 37°C and CO_2_ (5%). The diluted inoculum was then prepared in sterile 0.45% saline solution by resuspending the colonies until a turbid suspension equivalent to that of a McFarland 0.5 standard was obtained. Subsequently, following the protocols established by the Clinical and Laboratory Standards Institute (42,43), the identification tests (GP Test Cards for Gram-positive cocci) and antimicrobial sensitivity (AST03 Cards for Streptococcal Susceptibility) were performed, using the VITEK system-2 from BioMérieux. In short, the AST03 cards were inoculated, filled and inserted into the VITEK-2 incubator reader device within 15 minutes after the preparation of the inoculum, according to the manufacturer’s instructions. The AST03 susceptibility cards of the VITEK-2 system contain Wilkins-Chalgren culture medium, modified with the following antimicrobial agents: Benzyl-penicillin (Meningitis, Oral and Pneumonia), Ceftriaxone (Meningitis and Other), Cefotaxime (Meningitis and Other), Vancomycin, Erythromycin, Tetracycline, Clindamycin, Chloramphenicol, Linezolid, Tigecycline, Trimethoprim / Sulfamethoxazole, Levofloxacin, Moxifloxacin, Rifampicin. The strain ATCC 49619 was used as a pneumococcal control. According to the CLSI criteria, colonizing pneumococcal isolates were classified as sensitive, sensitive intermediate or resistant, according to established cut-off points (2,42,43).

### 2.7. Statistical Analysis

The statistical analysis of the data and results obtained in this study was performed as follows: 1) The data were tabulated, validated and analyzed using the Excel® program and the Statistical Package for Social Sciences (SPSS®) version 20.0 (IBM Corporation, Chicago, IL, USA); 2) Descriptive statistics (Univariate Analysis) were performed to summarize the sociodemographic information, the proportions of nasopharyngeal carriage, the molecular characteristics and susceptibility / antimicrobial resistance of the isolates; 3) Fisher and Chi-square exact tests (Bivariate Analysis) were carried out to identify the candidate variables for the multivariate analysis (Hosmer-Lemeshow Criteria, Cut Point P <0.25); and finally, 4) a logistic regression model (Multivariate Analysis) was made, using the Enter method, in order to identify possible factors associated with the nasopharyngeal carriage of pneumococci. P values <0.05 were considered statistically significant.

## 3. Results

### 3.1. Socio-Demographic, Clinical, Housing and Life Habits Characteristics

A total of 452 individuals under 5 years old of southwestern Colombia, who attended the Club Noel Children’s Clinic for outpatient services between September and October of 2019, were included in this study. From this population, 241 (53.3%) were male and 211 (46.7%) were female. According to the age, 105 (23.2%) children were under 1 year old, 85 (18.8%) were one year old, 88 (19.5%) children were two years old, 77 (17.0%) were three years old, 69 (15.3 %) four years, and 28 (6.2%) five years old. The average age of the children was 30 months with a standard deviation of 18 months. Regarding race or ethnicity, 349 (77.2%) identified themselves as mestizo-Colombians, 68 (15.1%) declared themselves to be Afro-Colombians, and 35 (7.7%) recognized themselves as indigenous people belonging to Amerindian communities settled mainly in the department from Cauca. The majority of children 429 (94.5%) belonged to socioeconomic strata 1, 2 and 3 (Low/medium-income levels), while two (0.4%) participants belonged to stratum 6 (high-income level) from the municipalities of Yumbo and Jamundí. Three hundred ninety-nine (88.3%) participating children came from 21 different municipalities in the department of Valle del Cauca of which 244 (61.1%) lived in the city of Cali. Participation of 51 (11.3%) children was obtained from the department of Cauca, while from the department of Nariño no children could be included during the sampling period (Table 1A).

**Table 1.** Sociodemographic (A), clinics (B) and housing and lifestyle habits (C) characteristics of the 452 outpatient children of the Club Noel Children’s Clinic, involved in the study, in the year 2019.

| **A. Sociodemographic Characteristics** |  |  |  |
| --- | --- | --- | --- |
| **Variable** |  | **n** | **%** |
| Gender | Male | 241 | 53,3 |
|  | Female | 211 | 46,7 |
| Age in years | 0 | 105 | 23,2 |
|  | 1 | 85 | 18,8 |
|  | 2 | 88 | 19,5 |
|  | 3 | 77 | 17,0 |
|  | 4 | 69 | 15,3 |
|  | 5 | 28 | 6,2 |
| Department | Valle del Cauca | 399 | 88,3 |
|  | Cauca | 2 | 0,4 |
|  | Nariño | 0 | 0,0 |
|  | Putumayo | 51 | 11,3 |
| Race or Culture | Native | 35 | 7,7 |
|  | Afro-Colombian | 68 | 15,1 |
|  | Mestizo-Colombian | 349 | 77,2 |
| Socioeconomic Stratum | 1 | 132 | 29,2 |
|  | 2 | 184 | 40,7 |
|  | 3 | 113 | 25,0 |
|  | 4 | 16 | 3,5 |
|  | 5 | 5 | 1,2 |
|  | 6 | 2 | 0,4 |
| **B. Clinical Characteristics** |  |  |  |
| **Variable** |  | **n** | **%** |
| Severe Underlying Disease (Total) | Yes | 42 | 9,3 |
|  | No | 410 | 90,7 |
| Previous Respiratory Diseases (Total) | Yes | 174 | 38,5 |
|  | No | 278 | 61,5 |
| Previous Respiratory Disease (Asthma) | Yes | 48 | 10,6 |
|  | No | 404 | 89,4 |
| Previous Respiratory Disease (Tonsillitis) | Yes | 24 | 5,3 |
|  | No | 428 | 94,7 |
| Previous Respiratory Disease (Bronchitis) | Yes | 39 | 8,6 |
|  | No | 413 | 91,4 |
| Previous Respiratory Disease (Laryngitis) | Yes | 13 | 2,9 |
|  | No | 439 | 97,1 |
| Previous Respiratory Disease (Otitis) | Yes | 37 | 8,2 |
|  | No | 415 | 91,8 |
| Previous Respiratory Disease (Rhinitis) | Yes | 28 | 6,2 |
|  | No | 424 | 93,8 |
| Previous Respiratory Disease (Sinusitis) | Yes | 22 | 4,9 |
|  | No | 430 | 95,1 |
| Previous Respiratory Disease (Pneumonia) | Yes | 51 | 11,3 |
|  | No | 401 | 88,7 |
| Previous Respiratory Disease (Meningitis) | Yes | 4 | 0,9 |
|  | No | 448 | 99,1 |
| Respiratory Signs and Symptoms (Total) | Yes | 267 | 59,1 |
|  | No | 185 | 40,9 |
| Respiratory Signs and Symptoms (Fever) | Yes | 33 | 7,3 |
|  | No | 419 | 92,7 |
| Respiratory Signs and Symptoms (Nasal Secretion) | Yes | 169 | 37,4 |
|  | No | 283 | 62,6 |
| Respiratory Signs and Symptoms (Sneezing) | Yes | 127 | 28,1 |
|  | No | 325 | 71,9 |
| Respiratory Signs and Symptoms (Cough) | Yes | 166 | 36,7 |
|  | No | 286 | 63,3 |
| Respiratory Signs and Symptoms (Phlegm) | Yes | 91 | 20,1 |
|  | No | 361 | 79,9 |
| Respiratory Signs and Symptoms (Difficulty Breathing) | Yes | 20 | 4,4 |
|  | No | 432 | 95,6 |
| Respiratory Signs and Symptoms (Fatigue) | Yes | 7 | 1,5 |
|  | No | 445 | 98,5 |
| Low Weight at Birth | Yes | 101 | 22,3 |
|  | No | 351 | 77,7 |
| Vaccination Status | Non-Immunized | 154 | 34,1 |
|  | Incomplete Immunization | 36 | 8,0 |
|  | Immunization in Process | 73 | 16,1 |
|  | Complete Immunization | 187 | 41,8 |

C. Characteristics of the Living Quarters, Home and Life Habits
| Variable |  | n | % |
| --- | --- | --- | --- |
| Overcrowding at Home | Yes | 51 | 11,3 |
|  | No | 401 | 88,7 |
| Shared Bedroom | Yes | 354 | 78,3 |
|  | No | 98 | 21,7 |
| Living with Smokers | Yes | 61 | 13,5 |
|  | No | 391 | 86,5 |
| Contact With Sick Person(s) (Otitis) | Yes | 18 | 4,0 |
|  | No | 434 | 96,0 |
| Contact With Sick Person(s) (Sinusitis) | Yes | 22 | 4,9 |
|  | No | 430 | 95,1 |
| Contact With Sick Person(s) (Bronchitis) | Yes | 7 | 1,5 |
|  | No | 445 | 98,5 |
| Contact With Sick Person(s) (Pneumonia) | Yes | 5 | 1,1 |
|  | No | 447 | 98,9 |
| Contact With Sick Person(s) (Meningitis) | Yes | 2 | 0,4 |
|  | No | 450 | 99,6 |
| Hospitalized Home Members | Yes | 27 | 6,0 |
|  | No | 425 | 94,0 |
| Assists to Child Care Center | Yes | 178 | 39,4 |
|  | No | 274 | 60,6 |
| **Total** |  | **452** | **100** |

Forty-two (9.3%) participating children were clinically diagnosed with severe diseases such as anemia, cerebral paralysis and hip dysplasia, among others. At some point in their life, 174 (38.5%) children were diagnosed with a respiratory illness such as pneumonia, asthma, bronchitis, otitis and rhinitis, among others. However, they were fully recovered at the time of sampling. On the other hand, 267 (59.1%) participants were diagnosed with respiratory signs and symptoms at the time the samples were collected, such as nasal secretion, cough, sneezing and phlegm, among others. Regarding the immunization status, 298 (65.9%) participants had received at least one dose of pneumococcal vaccine, mainly from the biological 10-valent conjugate (99.7%). Only 187 (41.4%) participants certified to have a complete PCV immunization schedule (2+1 doses) (Table 1B).

Regarding the conditions of housing, home and life habits, 51 (11.3%) children participating in the study lived in overcrowded conditions (3 or more people per bedroom), while 98 (21.7%) children had their own room and slept without companions in their own bed. Finally, 61 (13.5%) participants lived with people who smoke cigarettes regularly, while 178 (39.4%) of the children attended child care institutions near their homes (Table 1C).

### 3.2. Nasopharyngeal carriage of *S. pneumoniae* in Children of the Colombian Southwest

Of the 452 children examined, 189 (41.8%) were carriers of *S. pneumoniae*. The highest proportion of nasopharyngeal colonization of *S. pneumoniae* was observed in two-year-old children (41 children, 46.6%). The overall proportion of nasopharyngeal carriage of *S. pneumoniae* was 105 (43.6%) in males versus 84 (39.8%) in females. Children belonging to Indigenous communities had the highest proportion of nasopharyngeal colonization (22 children, 62.9%), while the lowest proportion was observed in Afro-Colombian participants (24 children, 35.3%). In children with the lowest socioeconomic condition (Stratum 1), the overall proportion of nasopharyngeal carriage of *S. pneumoniae* was 63 (47.7%). The colonization rate of children from the department of the Cauca was 58.8% (Table 2A).

**Table 2.**
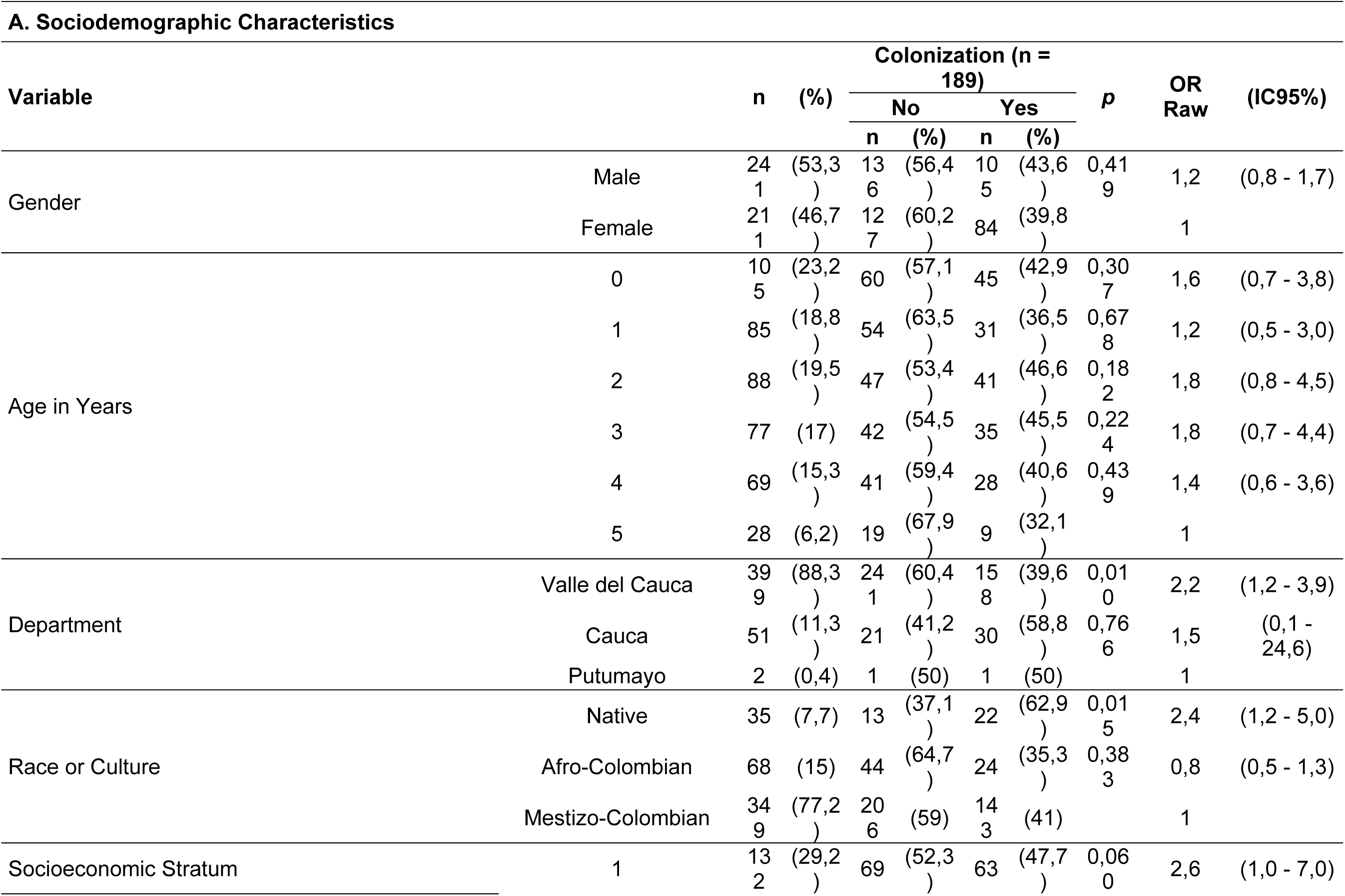
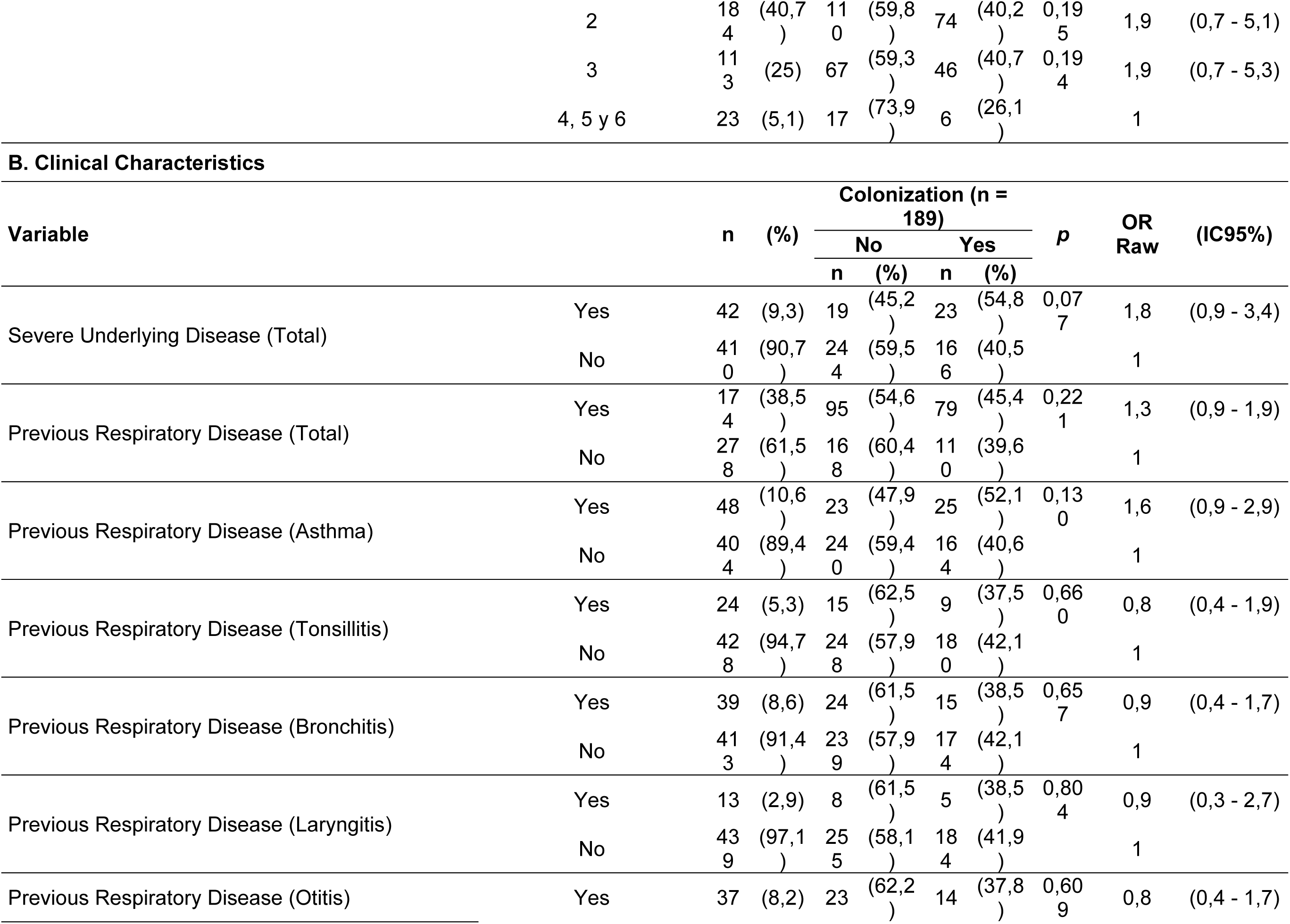
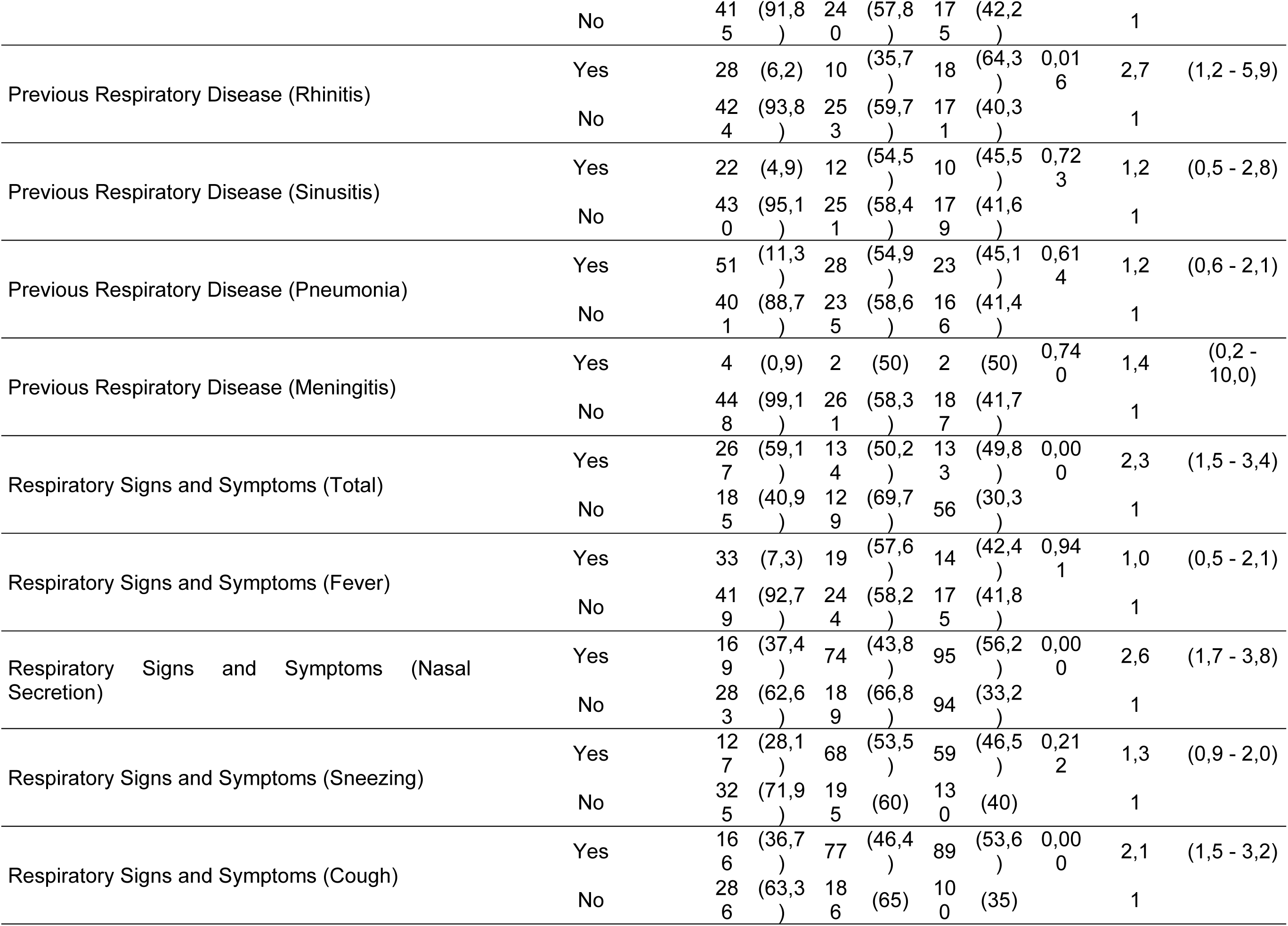
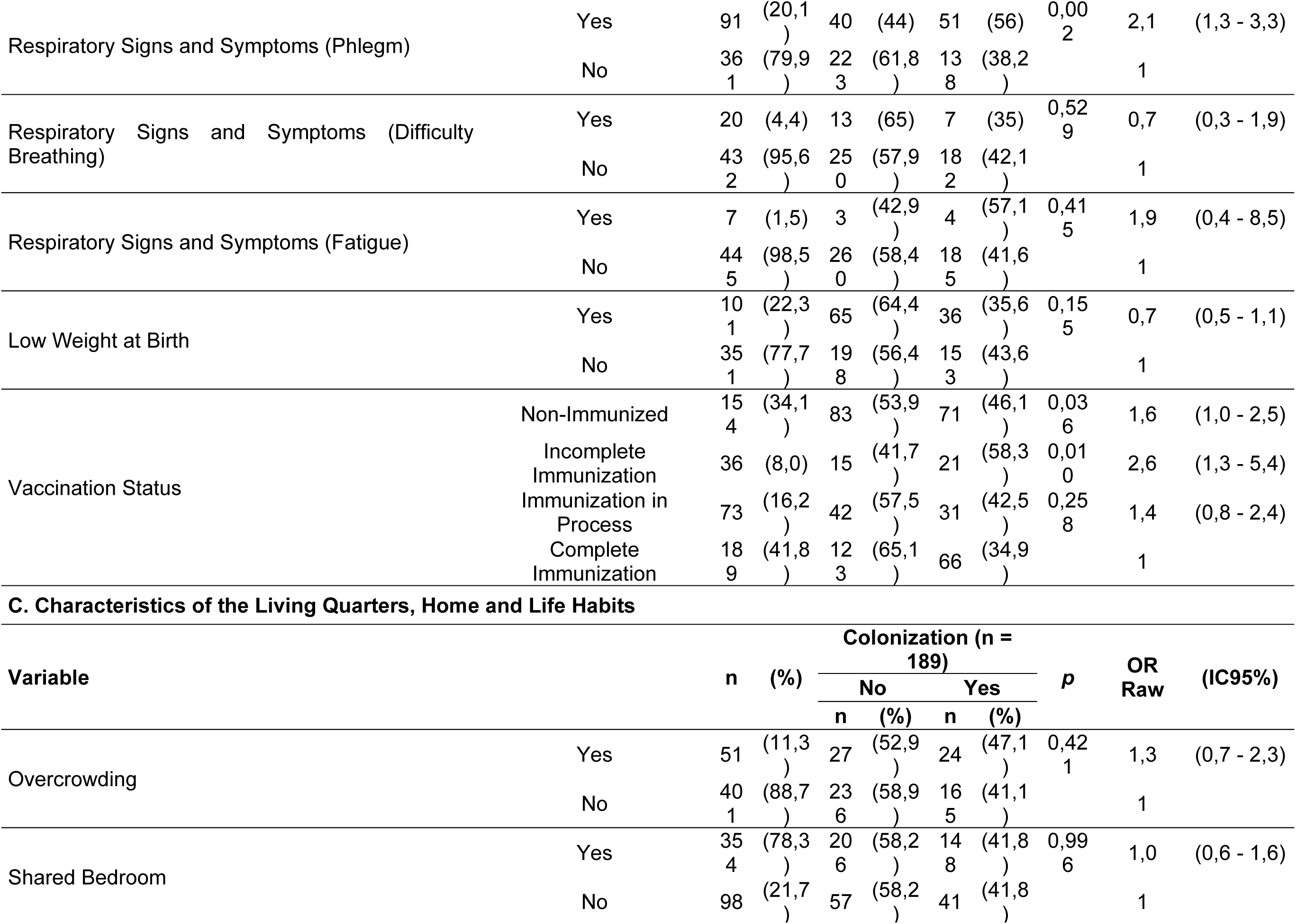
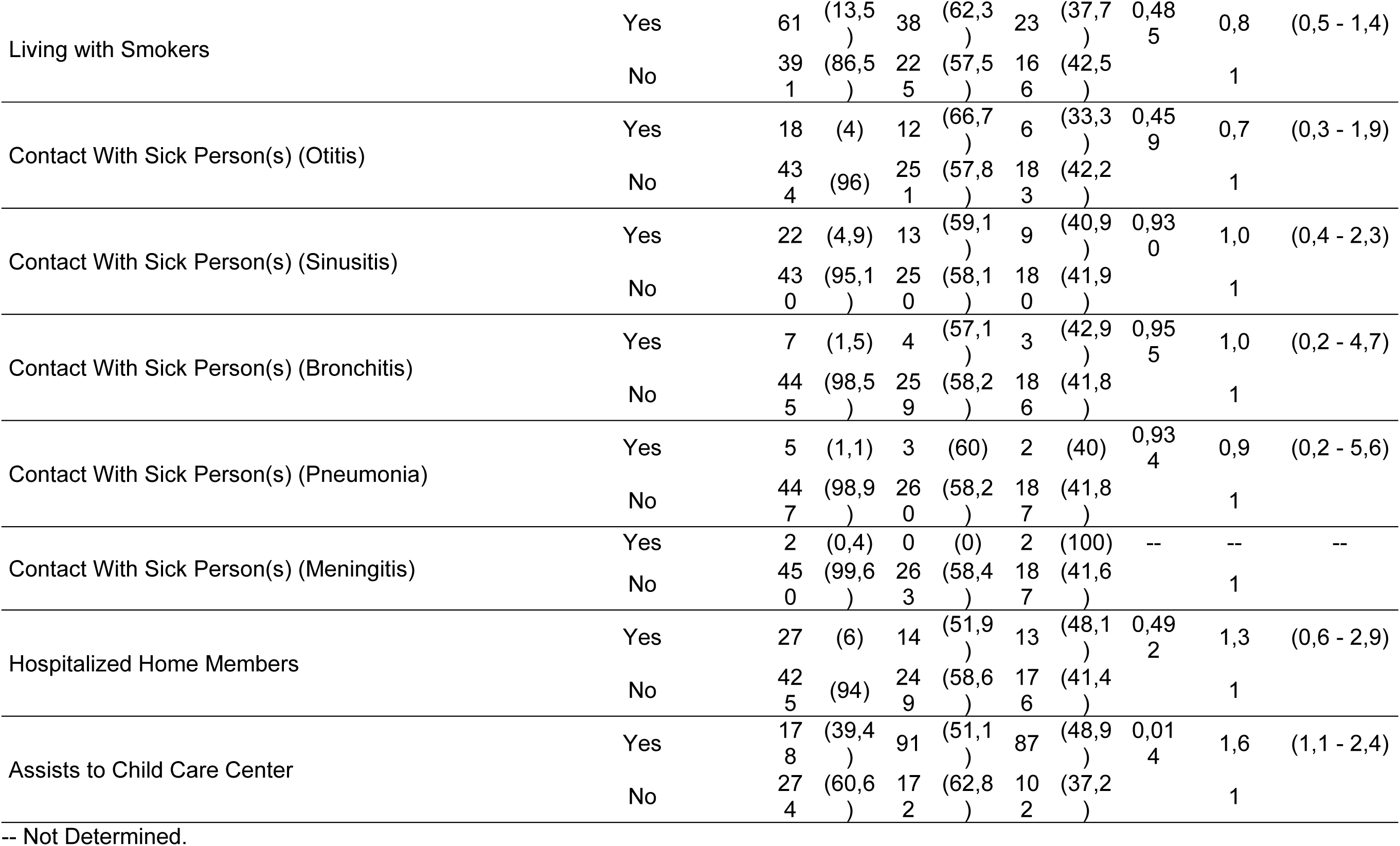
Bivariate analysis of associated factors for *Streptococcus pneumoniae* nasopharyngeal colonization in 452 outpatient children of the Club Noel Children’s Clinic, involved in the study, in 2019.

Having been clinically diagnosed with severe diseases (23, 54.8%), respiratory diseases at some time in life (79, 45.4%), and respiratory signs and symptoms at the time of sampling (133, 49.8%) were the variables identified with higher proportions of nasopharyngeal colonization by *S. pneumoniae* than in healthy participants. Likewise, the colonization ratios of children not immunized (71, 46.1%) or with incomplete immunization schedules (53, 47.7%) were higher than those children with complete immunization schedule (65, 34.8%) (Table 2B, Figure 1).

The proportion of nasopharyngeal colonization of *S. pneumoniae* was 24 (47.1%) among participants living in overcrowded conditions versus 41 (41.8%) in children who do not share their room and sleeping alone in their bed. Of the participants living with people smoking regularly at home, 23 (37.7%) were positive for *S. pneumoniae*. Finally, the proportion of colonization of children attending child care institutions in the vicinity of their homes (87, 48.9%) was higher than that of participants not attending any institution (102, 37.2%) (Table 2C).

### 3.3. Analysis of the Factors Associated with the Nasopharyngeal Carriage of *S. pneumoniae*

The results showed a correlation between pneumococcal colonization and the 2-year age group (OR = 3.0; 95% CI = 1.0-8.3; p = 0.041). The nasopharyngeal carriage of *S. pneumoniae* was significantly higher in children belonging to indigenous communities (OR = 2.7; 95% CI = 1.2-5.9; p = 0.014). In addition, there was a significant association between nasopharyngeal colonization of *S. pneumoniae* and nasal secretion at the time of sampling (OR = 2.1; 95% CI = 1.3-3.3; p = 0.003). Failure to complete immunization schedules (OR = 2.5; 95% CI = 1.3-5.0; p = 0.008) and not having received immunization against pneumococcus (OR = 1.7; 95% CI = 1.1-2.8; p = 0.028) was significantly associated with the presence of *S. pneumoniae* in the nasopharynx in southwestern Colombia (Figure 2). Likewise, attending childcare institutions (OR = 1.8; 95% CI = 1.0-3.2; p = 0.042) was significantly correlated with nasopharyngeal colonization of *S. pneumoniae*. However, there was no significant association between gender, socioeconomic stratum, severe diseases, respiratory disease at some time in life (asthma, pneumonia, rhinitis, etc.), respiratory signs and symptoms at the time of sample collection (cough, sneezing and phlegm, among others), low weight at birth, and overcrowding with the nasopharyngeal carriage of *S. pneumoniae* (Table 3).

**Table 3.**
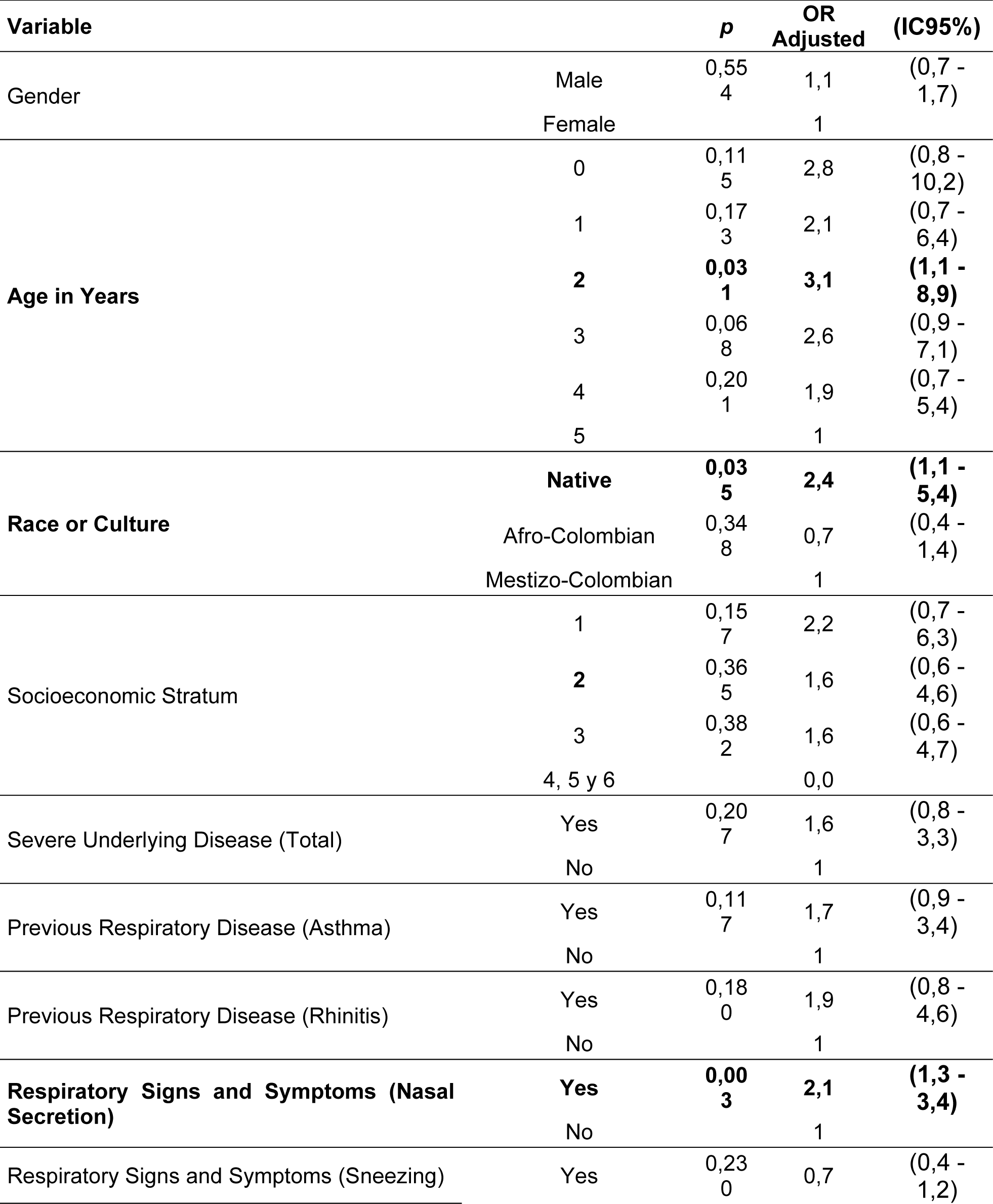
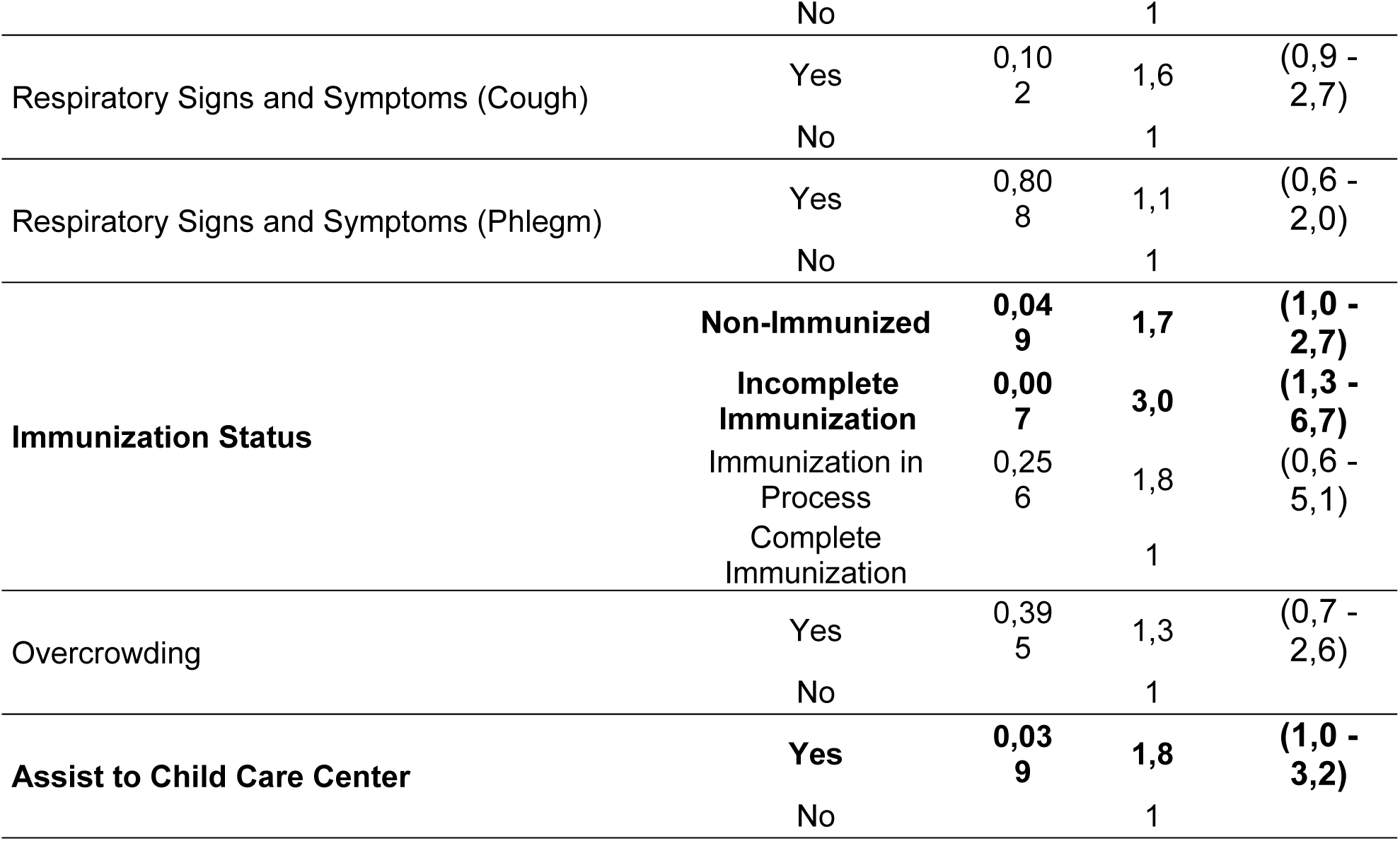
Multivariate analysis of associated factors for *Streptococcus pneumoniae* nasopharyngeal colonization in 452 outpatient children of the Club Noel Children’s Clinic, involved in the study, in 2019.

**Figure 2.**
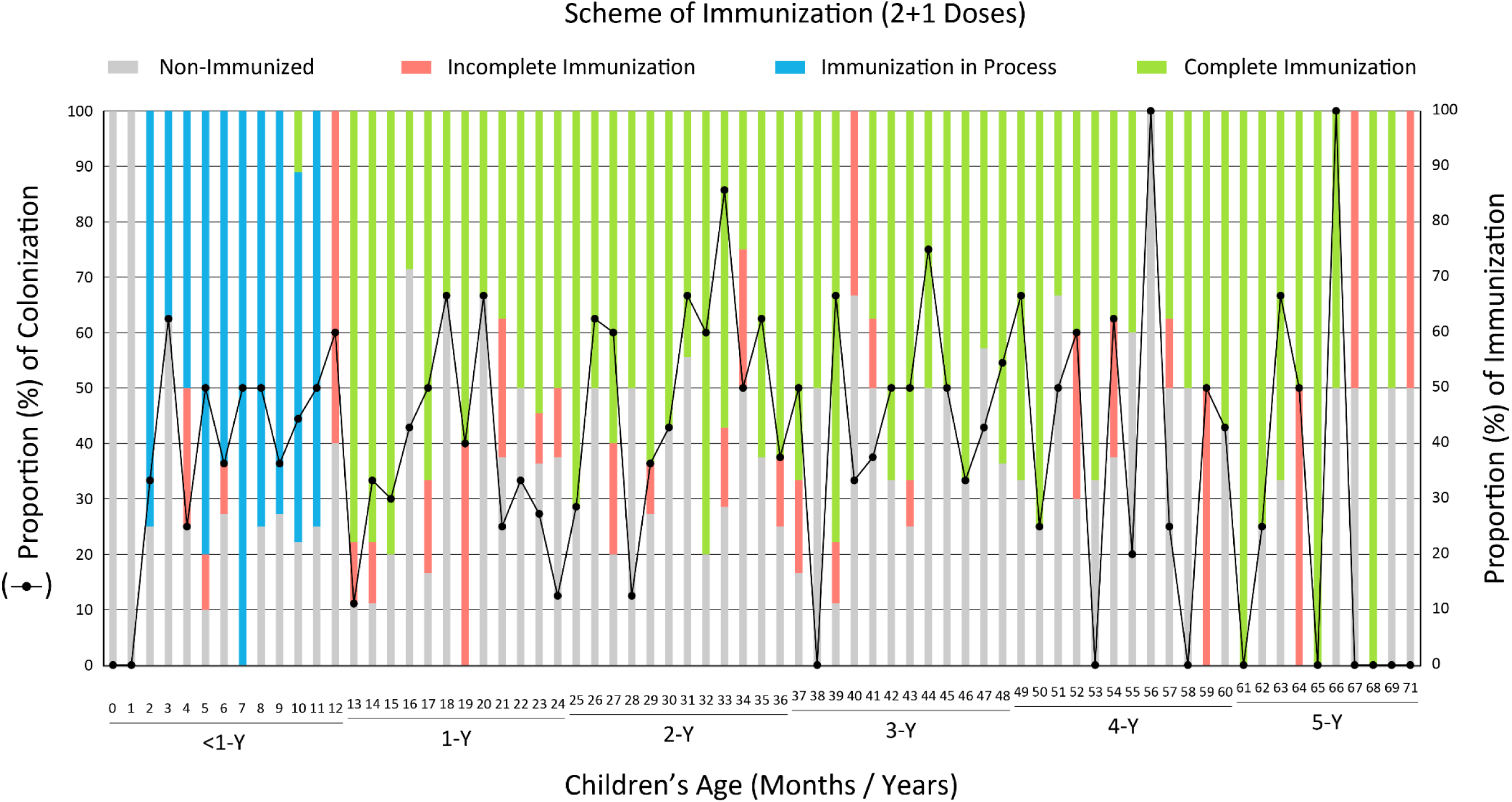
Proportion of nasopharyngeal colonization by *Streptococcus pneumoniae* and immunization status according to the age in months (and years) of participating children. Southwestern of Colombia, 2019.

### 3.4. Antimicrobial Resistance / Susceptibility Profiles of Colonizing Isolations

The resistance / susceptibility profiles to the 18 antibiotics contained in the AST-03 card (VITEK-2) of the *S. pneumoniae* colonizing isolates identified in this study are reported in Table 4. All pneumococcal isolates were susceptible to vancomycin, chloramphenicol, linezolid, tigecycline, levofloxacin, moxifloxacin and rifampicin. Fifty-five (29.1%) *S. pneumoniae* colonizing isolates were susceptible to all antibiotics tested, 31 (16.4%) were resistant to an antimicrobial agent, and 26 (13.8%) were resistant to two. Sixty-nine (36.5%) colonizing isolates of *S. pneumoniae* were resistant to between three and ten different antibiotics (MDR: Multi-Drug Resistant), while 23 (12.2%) presented resistance profiles to at least one antibiotic in each class of antimicrobial agents (XDR: Extensively-Drug Resistant). 109 (57.7%) *S. pneumoniae* clinical isolates not susceptible to benzyl-penicillin (cut meningitis) were identified and isolated, and 86 (45.5%) presented a reduced or intermediate susceptibility to this antibiotic, according to the oral cutoff threshold. Seventy-seven (40.7%) colonizing isolates of *S. pneumoniae* were resistant to erythromycin, 68 (36.0%) were resistant to tetracycline, 47 (24.9%) were resistant to trimethoprim / sulfamethoxazole, and 46 (24.3%) were resistant to clindamycin. Regarding Ceftriaxone and Cefotaxime (third generation cephalosporin), 51 (27.0%) and 41 (21.7%) isolates of *S. pneumoniae* were identified with resistance profiles and intermediate susceptibility, respectively.

**Table 4.** Antimicrobial susceptibility profiles of *Streptococcus pneumoniae* colonizing strain, isolates of the 452 patients involved in the study.

| **Antimicrobial Agent** | ***S. pneumoniae* (n = 189)** |  |  |  |  |  |
| --- | --- | --- | --- | --- | --- | --- |
|  | **Resistant** |  | **Intermediate** |  | **Susceptible** |  |
|  | **n** | **(%)** | **n** | **(%)** | **n** | **(%)** |
| Benzyl-penicillin (Meningitis) | 109 | (57,7) | 0 | (0) | 80 | (42,3) |
| Erythromycin | 77 | (40,7) | 0 | (0) | 112 | (59,3) |
| Tetracycline | 68 | (36,0) | 0 | (0) | 121 | (64,0) |
| Trimethoprim/sulfamethoxazole | 47 | (24,9) | 15 | (7,9) | 127 | (67,2) |
| Clindamycin | 46 | (24,3) | 5 | (2,6) | 138 | (73,0) |
| Benzyl-penicillin (Oral) | 23 | (12,2) | 86 | (45,5) | 80 | (42,3) |
| Ceftriaxone (Meningitis) | 17 | (9,0) | 14 | (7,4) | 158 | (83,6) |
| Cefotaxime (Meningitis) | 15 | (7,9) | 14 | (7,4) | 160 | (84,7) |
| Ceftriaxone (Another) | 10 | (5,3) | 7 | (3,7) | 172 | (91,0) |
| Cefotaxime (Another) | 9 | (4,8) | 6 | (3,2) | 174 | (92,1) |
| Benzyl-penicillin (Pneumonia) | 4 | (2,1) | 5 | (2,6) | 180 | (95,2) |
| Vancomycin | 0 | (0) | 0 | (0) | 189 | (100) |
| Chloramphenicol | 0 | (0) | 0 | (0) | 189 | (100) |
| Linezolid | 0 | (0) | 0 | (0) | 189 | (100) |
| Tigecycline | 0 | (0) | 0 | (0) | 189 | (100) |
| Levofloxacin | 0 | (0) | 0 | (0) | 189 | (100) |
| Moxifloxacin | 0 | (0) | 0 | (0) | 189 | (100) |
| Rifampicin | 0 | (0) | 0 | (0) | 189 | (100) |

## Discussion

In this study, a nasopharyngeal carriage rate of *S. pneumoniae* in children under 5 years of 41.8% was observed for the first time in Colombia. These results are in accordance with the high frequencies of colonization observed by our research group in other cities of the country, such as Medellín, where it has been possible to observe a colonization of 55.5% (unpublished data). Other surveillance studies conducted in other countries have reported similar carriage rates (17,35,37,44). This may, nevertheless, suggest that the Colombian southwest has a large number of children belonging to low socioeconomic strata, carriers of *S. pneumoniae*, with a high risk of developing invasive and non-invasive pneumococcal diseases. Consequently, they represent a transmission reservoir, not only for members of their families (other children and older adults), but also for their childhood partners with whom they interact in the community, especially in institutions where care is provided (45). In our study, children belonging to the age group of 2 years were identified with highest carriage rate (41: 46.6%), which coincides with the times when parents begin their children’s schooling in this region of the country. Likewise, participants who said they belonged to indigenous communities were the most vulnerable to the nasopharyngeal presence of *S. pneumoniae* (22: 62.9%), which also contributed to a colonization rate well above the average for the department of Cauca (30: 58.8%). These results coincide with reports from countries with similar conditions to those in this Latin American region (23,46–50). However, in children 3, 4 and 5 years of age, a non-significant decrease in the proportion of colonization by *S. pneumoniae* was observed, which could be the reflection of the gradual acquisition of mucosal immunity from the respiratory tract superior, especially in children with complete conjugate immunization schemes against pneumococcus, and reduced exposure to the pathogen in nurseries and their homes. These findings confirm that children under 2 years of age have a higher risk of acquiring *S. pneumoniae* and suffering from their diseases, compared to the older age groups (3, 4 and 5 years).

Various studies worldwide have described several sociodemographic characteristics associated with an increase in nasopharyngeal carrying of *S. pneumoniae*, which includes low birth weight, overcrowding, sleeping with parents or other family members, and the fact of living with people who smoke regularly (15). However, in our study, none of these factors was associated with nasopharyngeal colonization of *S. pneumoniae*. On the contrary, other factors such as environmental and / or host factors (underlying diseases, immunosuppression, etc.) could be the main determinants of the distribution of the carriage among the children of the Colombian southwest participating in this study. In addition to age and ethnicity, in the logistic regression analysis, colonization rates for *S. pneumoniae* were significantly associated with nasal secretion at the time of sampling, and with not being immunized against pneumococcus or having the incomplete immunization schedule (2 + 1) (Figure 2) (3,18). Regarding the environmental factor, children attending childcare institutions showed a significantly greater nasopharyngeal colonization of *S. pneumoniae* compared to children who were not in school. These results are similar to those found by our research group in children attending school in the city of Medellín and to reports from countries with similar conditions (51–53). This finding is of great relevance because exposure to other children during childhood, especially peers in community care institutions, has been clearly associated with an increased risk of colonization and invasive and non-invasive pneumococcal disease.

The results of the susceptibility study revealed higher antibiotic resistance of *S. pneumoniae* to relatively cheap and readily available antibiotics for the population such as benzyl-penicillin cut meningitis (57.7%), erythromycin (40.7%), tetracycline (36.0%) and trimethoprim/sulfamethoxazole (24.9%), and more expensive but of variable use alternatives such as ceftriaxone (27.0%) and clindamycin (24.3%). This observation is consistent with previous reports in Venezuela and other countries of the world (33,54–58). On the other hand, vancomycin, chloramphenicol, linezolid, tigecycline, levofloxacin, moxifloxacin and rifampicin were the most effective antibiotics against *S. pneumoniae* isolates, all with 100% antimicrobial susceptibility, which is consistent with reports from other countries of the region (30,33,59). Twenty-three (12.2%) colonizing isolates of *S. pneumoniae* presented resistance profiles to at least one antibiotic in each class of antimicrobial agents, which includes them in the dangerous group of extensively drug-resistant pneumococci (XDR). Likewise, 69 (36.5%) isolates were resistant to between three and ten different antibiotics, being considered in this study as multi-drug resistant *S. pneumoniae* (MDR). Less than 1/3 of the pneumococci isolated in southwestern Colombia were susceptible to all antibiotics tested, which is a direct product of frequent and inappropriate use of the chemotherapeutics. Although data on the use of different antibiotics in low-and middle-income countries are underrepresented, the rates of resistance of pneumococci to antimicrobial agents vary according to geographic region and the different population subgroups analyzed (31,34,60). These variations represent major challenges for health systems in Latin American countries and reflect the uncontrolled and low-cost availability of some of these medical resources (30). This phenomenon is not foreign to Colombia and its southwestern region (14,61–63), which would be exerting greater selection pressure for resistant *S. pneumoniae* strains, favoring the increase in their frequency and, therefore, decreasing the efficacy of these antibiotics in the treatment of *S. pneumoniae* affected patients.

## Conclusions

In this study, a general proportion of nasopharyngeal colonization of *S. pneumoniae* of 41.8% is reported for the southwestern region of Colombia, with a higher frequency among 2-year-old participants. Belonging to Native American (indigenous) communities, not being immunized against pneumococcus, not completing established immunization schemes, attending child-care institutions, and presenting nasal secretion are risk factors for nasopharyngeal carrying of *S. pneumoniae* in this region of the country. On the other hand, a non-susceptibility of *S. pneumoniae* to benzyl-penicillin (meningitis and oral cuts), increased resistance to antibiotics erythromycin, tetracycline, trimethoprim/sulfamethoxazole and clindamycin was observed, in addition to resistance and intermediate levels of susceptibility to cephalosporin of broad spectrum (ceftriaxone and cefotaxime). In conclusion, with this study the local and regional frequency data of children under 5 years of age carrying *S. pneumoniae* is obtained for the southwest of Colombia for the first time. This high proportion of children carrying *S. pneumoniae* could show an important reservoir of bacterial transmission among children under 5 years of age in that community, which could potentially lead to the onset of pneumococcal diseases with serious consequences for the health of people in this Colombian region. Therefore, there is a clear need to expand pneumococcal conjugate immunization in the community and ensure compliance with established immunization schedules. Additionally, the determination of the association of nasopharyngeal colonization of resistant MDR and XDR-like strains with the development of invasive infection by resistant strains is important to establish rational treatments for the alleged *S. pneumoniae* infections in southwestern Colombia.

## Data Availability

Detailed information will be provided after a reasonable request.

## Acknowledgements

The authors thank Jaime Dominguez Navia, Luz Myriam Claros, María Victoria Hernández, María Victoria Muñoz, María del Palmar, Jhonny Castrillón, and all the staff of Club Noel Children’s Clinical Foundation, Cali, Colombia for facilitating the realization of this study.

## Authors’ Contribution

Conceptualization: GG, JPR, LFM, JCO, LJA, JAB, AGM, JLM, SH.

Data curation: GG, JPR, SC, MAP, JLT, LMM, LFV, YAZ, JLM.

Formal analysis: GG, JPR, JLM, SH.

Funding acquisition: GG.

Investigation: GG, JPR, SC, JDC, MAP, LFM, JLT, JCO, LMM, JCR, LFV, AMF, YAZ, MEC, LJA, AJM, JAB, AGM, SH.

Methodology: GG, JPR, SC, JDC, MAP, LFM, JLT, JCO, LMM, JCR, LFV, AMF, YAZ, MEC, LJA, AJM, JAB, AGM, SH.

Project administration: GG.

Resources: GG.

Software: Not applicable.

Supervision: GG, JPR, SH.

Validation: GG, JLM.

Visualization: GG, JLM.

Writing-original draft: GG, AGM, JLM.

Writing-review & editing: GG, JPR, SC, JDC, MAP, LFM, JLT, JCO, LMM, JCR, LFV, AMF, YAZ, MEC, LJA, AJM, JAB, AGM, JLM, SH.

